# Physical activity, sedentary behaviour, and sleep on Twitter: A labelled dataset for public health research

**DOI:** 10.1101/2021.04.13.21255449

**Authors:** Zahra Shakeri Hossein Abad, Gregory P. Butler, Wendy Thompson, Joon Lee

**Author notes:** corresponding author(s): Zahra Shakeri Hossein Abad.

## Abstract

Advances in automated data processing, together with the unprecedented growth in user-generated social media (SM) content, have made public health surveillance (PHS) one of the long-lasting SM applications. However, the existing PHS systems feeding on SM data have not been widely deployed in national surveillance systems, which appears to stem from the lack of practitioners’ trust in SM data. More robust datasets over which machine learning (ML) models can be trained/tested reliably is a significant step toward overcoming this hurdle. The health implications of physical activity, sedentary behaviour, and sleep (PASS) are widely studied through traditional data sources, which are often out-of-date, costly to collect, and thus limited in quantity and coverage. We present LPHEADA, a multicountry and fully Labelled digital Public HEAlth DAtaset of tweets originated in Australia/Canada/United Kingdom/United States between November 2018-June 2020. LPHEADA contains 366,405 labels for 122,135 PASS-related tweets and provides details about the place/time/demographics associated with each tweet. LPHEADA is publicly available and can be utilized to develop (un)supervised ML models for digital PASS surveillance.

## Background & Summary

Almost two-thirds of the world’s population now use the Internet, taking the global total to 4.57 billion (59%) by July 2020^1^. 87% of Internet users and 65% (3.96 billion) of the world’s total eligible population (i.e. over the age of 13) now use social media. The combined time that these users spend on social media adds up to more than 1 million years every day^1^, contributing to a large amount of user-generated data on different social media platforms. In 2020, Twitter alone reported 500 million tweets generated per day from 145 million daily active users. The low-cost data stream available on social media and other Internet-based sources serves to make research advances on Digital Public Health Surveillance (DPHS) more accessible for public health officials, clinicians, patients, and the general public. This help to disseminate insights into different aspects of public health and promote healthy lifestyles and health policies^2,3^. The open access to the public data about users and their opinions, the ease of use, and a large user base have made Twitter one of the most popular data sources for studying different aspects of public health^4,5^, with Google Scholar indexing 1.32 million articles mentioning Twitter and public health. Moreover, more than 85% of Twitter users, with a wide breadth of demographic groups^4^, also use Facebook, Instagram, and YouTube (this number for other platforms varies between 52% to 82%)^1^, indicating that Twitter users reasonably represent active social media users in general.

Since 2011, Twitter has been the most popular form of social media used for public health communication^6,7^. A recent scoping review of 755 articles on DPHS shows that Twitter is the most studied of all platforms and most utilized platform to study communicable diseases, behavioural risk factors, mental health, drug utilization, and vaccine^7^.

However, a number of limitations that mainly stem from the limitations associated with the data, are still the major obstacles towards the adoption of digital data for public health surveillance^4,7^. Given the main aims of any PHS system are to measure, monitor, and improve the overall health status of their target populations, the systematic incorporation of time, demographics (i.e. age, gender), and place data into the surveillance process is critical to the reliability and generalizability of this process^8,9^. However, nearly one-third (32%) of the digital public health surveillance studies published between 2005-2020 (with the majority of them related to behavioural risk factors surveillance) did not capture age, gender, or place information for their analyses^7^. Moreover, most studies on DPHS do not consider whether their findings are associated with the user’s personal experience (self-reported or not), leading to content bias, incorrect results and potentially flawed interpretations^7^.

Considering the location-dependent nature of health policies, along with the essential role of place data in assessing the representativeness of a PHS system, the impact of a PHS system can vary considerably with geographical location^10–13^. However, the number of DPHS studies that have stratified their results by more granular geographic region is small^7^. Due to a lack of annotated datasets for the development of automatic models, more than two-thirds (69%) of DPHS studies published before 2020 are limited by labour-intensive, manual, and abstract analysis methods (e.g. manual coding, qualitative analysis, and rule-based natural language processing), which makes these studies limited in terms of sample size, scope and generalizability^7^.

Given that all of these challenges are data-oriented, an increase in both data quality and quantity enriched with concrete demographics and location information can help deal with all these challenges. Moreover, to facilitate the development and evaluation of robust machine learning models to address the limited scope of manual data analysis techniques, annotated datasets for various PHS aspects are required. However, only a handful of annotated datasets are publicly available for research on DPHS^14–21^. Jimeno-Yepes et al.,^15^ provided an annotated dataset of 1,300 tweets related to disease symptoms and pharmacologic substances. The open dataset developed by Aphinyanaphongs et al.,^16^ contains 13,146 labelled tweets resulting from hashtag filtering and covers a time span from January 2010 to January 2015. This dataset is developed for training binary classifiers to detect tweets that indicate e-cigarette use for smoking cessation. Crowdbreaks^18^, an open health tracking platform, crowdsources the labelling of vaccine sentiment and COVID-19 related tweets to the public. While the dataset provided by this system, compared to other open DPHS datasets, is in a better position in terms of size, it lacks demographics and geospatial data, and each tweet is labelled by only one annotator (without any control over their labelling quality).

Given that in addition to physical inactivity, as the leading risk factor for non-communicable diseases and premature death^22^, prolonged sedentary behaviour and inadequate sleep are also important risk factors for chronic disease^23^, this work presents a multicountry and fully Labelled digital Public HEAlth DAtaset (LPHEADA) of tweets related to physical activity, sedentary behaviour, and sleep (PASS) that originated in Australia, Canada, the United Kingdom (UK), or the United States (US). We selected these countries as they have some of the highest proportions of social media users in the world (Australia: 71%, Canada: 66%, UK: 66%, US: 69%, and World: 51%)^1^. LPHEADA comprises 366,405 labels, labelled by 708 unique annotators on Amazon Mechanical Turk (AMT), for 122,135 unique tweets generated by 72,749 unique users between November 28^*th*^ 2018 to June 19^*th*^ 2020. AMT is a software service operated by Amazon that allows users (*aka* requesters) to crowdsource work–broken into micro-tasks called human intelligence tasks (HITs), to a large number of workers who are compensated for each HIT completed^24^. As LPHEADA was collected and labelled in collaboration with the Public Health Agency of Canada (PHAC) to develop PASS indicators for the Canadian population, around 81% (98,722) of the tweets included in our dataset were collected from Canada. Tweets from the US and UK make up 8% of the dataset each (US: 10,193, UK: 9,154), and Australian tweets make up the remaining 3% (4,067) of the dataset. Along with the labelled tweets, we provide detailed information about users’ gender, age range, geospatial information, whether the tweet is self-reported or not, and whether it is posted by an organization. We evaluated the quality of the dataset and its labels using latent semantic analysis (LSA), linguistic analysis, machine learning models, and truth inference models. The dataset we provide in this paper can be utilized to develop (un)supervised machine learning models for digital PASS surveillance.

## Methods

### Collection and Preparation of the Dataset

We collected the data of this study from Twitter using the `Twitter livestream application programming interface` (API) between 28^*th*^ November 2018 to 19^*th*^ June 2020. The entire dataset (i.e. 1,902,980,841 tweets) was filtered for Canadian tweets potentially relevant to PASS. 103,911 tweets were selected from 22,729,110 collected Canadian tweets using keywords and regular expressions related to PASS categories. Each of these 103,911 tweets was labelled by three AMT workers, from which 98,722 tweets received three valid labels (i.e. multi/empty-labels are invalid and were rejected), with almost half of them related to physical activity. For the Canadian dataset, 610 unique workers participated in our data labelling tasks and completed 103,911 HITs, from which 5,189 HITs were removed as they did not receive three valid answers. The majority of these workers (87%, 530) completed less than 100 HITs each, among which 164 completed only one HIT each. Among the workers who completed more than 5,000 HITs, one worker completed 21,801 HITs, and three workers completed between 5,000 and 10,000 HITs.

In addition to the Canadian tweets, we filtered a random subset of the dataset for tweets that originated in the UK, USA, and Australia. This dataset spans the same data collection period as the Canadian dataset and contains 70,239 labels collected for 23,413 tweets (i.e. three labels per tweet). Adding the data from these countries will provide an important epidemiological diversity that can be utilized for implementing comparative studies and evaluating the generalizability of the PASS surveillance models trained on the Canadian dataset.

### Labelling Process

We implemented a pipeline to create the crowdsourcing tasks, referred to as HITs by AMT, post them on AMT, collect the labels through a quality check process, approve/reject the HITs, and store the results. To minimize noisy and low-quality data, we added a qualification requirement to our tasks and granted the labelling access to workers who had demonstrated a high degree of success in performing a wide range of HITs across AMT (i.e. master qualification). In addition to this, we added a simple qualification question to each HIT to detect spammers or irresponsible workers. Each HIT contained four questions, including the qualification question and was assigned to three workers. Through different iterations of data labelling, workers were paid from US$0.03 to US$0.05 after completing each HIT. We collected the labels for the 122,135 tweets used in this study through different iterations, from April 2019 to November 2020. We regularly checked the quality of submitted tasks to detect low-quality workers during each iteration and revoke their access to our tasks. Before the formal initiation of the process, we pilot tested the design, response time, and complexity of the HITs in two iterations and revised the workflow accordingly. To label the dataset provided in this paper, we utilized AMT as a crowdsourcing service and did not collect any personally identifiable information from the workers (participants) during the data labelling task. The experiments were carried out in accordance with relevant guidelines and the University of Calgary Conjoint Faculties Research Ethics Board’s (CFREB) regulations. We implemented the entire workflow in `python` and used `Boto3` `python` `SDK` to connect to and work with AMT.

### Time Adjustment

Twitter API returns the date and time that a tweet is published in UTC. To adjust this timezone base on each tweet’s location, we utilized the bounding box of coordinates, which enabled spatial mapping to tweets’ respective city locations and used `timezonefinder` in `python`. Given that daytime, month, and weekday can be influential factors in twitting about each of the PASS categories, and to better utilize the datetime data (`%a` `%b` `%d` `%H`:`%M`:`%S` `%Y`), we extracted a: weekday, b: month, and H: hour fields and stored them as separate features. Figure 1a shows the temporal distribution of tweets for each of the PASS categories in the Canadian dataset. Moreover, the stacked area charts presented in Figures 1b-1d detail the frequency of tweets for each of PASS categories for the top ten Canadian cities.

**Figure 1.**
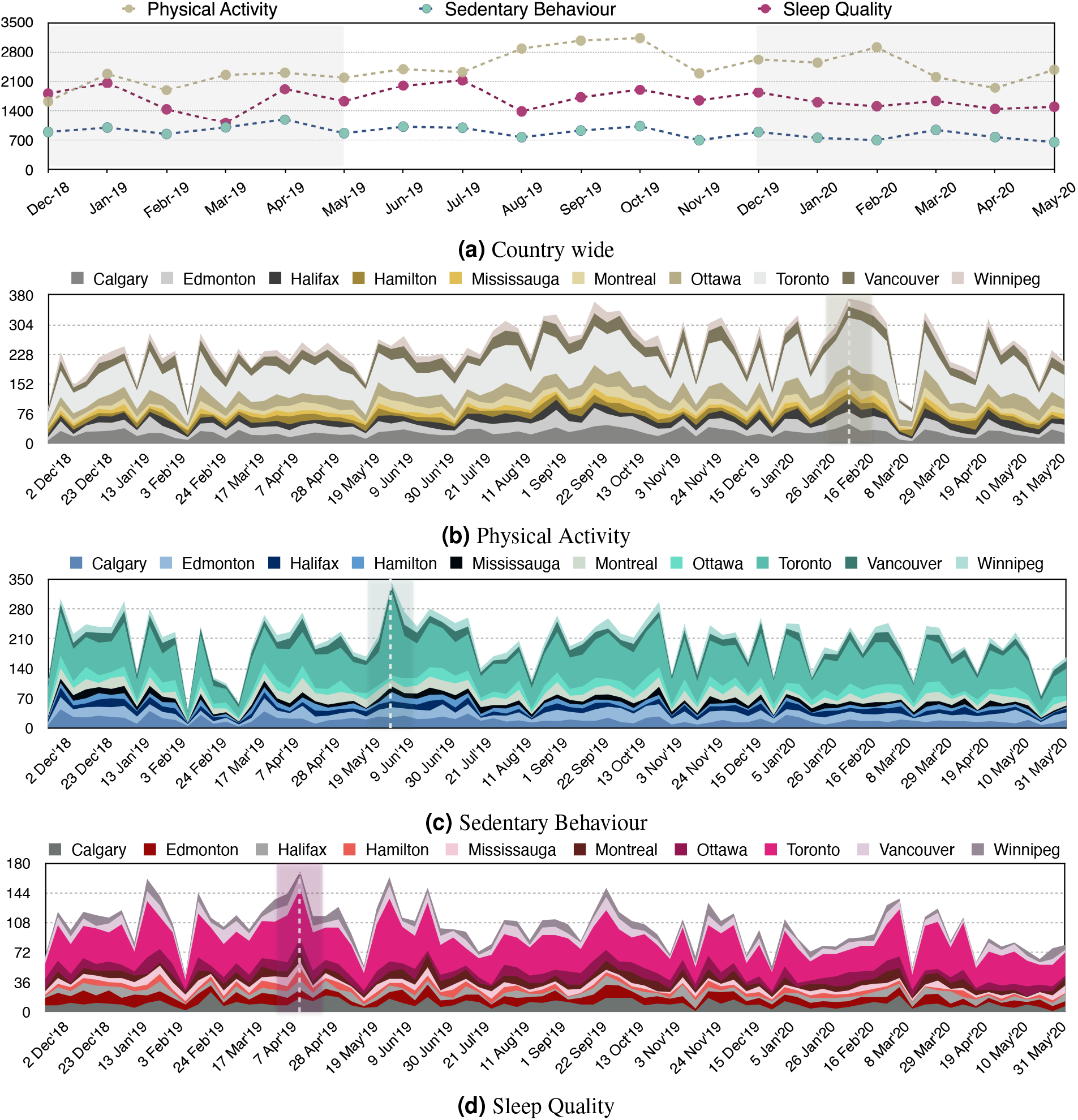
The temporal distribution of the tweets for the Canadian dataset. To make fair comparisons, we used the data from December 1^*st*^ 2018 to May 31^*st*^ 2020 for these visualizations and removed the data collected during the last two days of November 2018 and the first two weeks of June 2020.

### Location Inference

The geospatial metadata provided by the Twitter API is derived from three main sources, including the (1) geotagged location, (2) profile location, and (3) mentioned location in the tweet text. Geotagged location can be defined by exact location (i.e. device location) at the time of tweeting, by assigned twitter place (i.e. at the neighbourhood level), or both. While the exact location field provides the highest level of precision, it is not a default setting, and only a small portion of users share their exact latitude and longitude (e.g. only 1-2% of tweets are geo-tagged^25^). Thus, to infer the location of each tweet in LPHEADA, we proposed and developed a five-step process that utilizes tweet-neighbourhood location (i.e. place.name and place.full_name), profile information (i.e. profile description and location), and tweet text to map twitter’s geospatial metadata for each tweet *t*_*i*_ to physical locations in the form of ⟨*c*_*i*_, *p*_*i*_ | *s*_*i*_⟩, where *c* denotes the city of a tweet and *p* | *s* represent its corresponding province or state, respectively (Figure 2). To demo the proposed process, we used the Canadian geographical names dataset (i.e. location dictionary), provided by the Geographical Names Board of Canada. Each geographical name provided by this dataset is mapped to a province and is classified to a geographic area such as city, town, village, lake, administrative sectors, or recreational centers. As illustrated in Figure 2, for each *t*_*i*_, we first use a simple search function to map the ‘place.name’ field associated with each tweet to its corresponding *c*_*i*_ in the location dictionary (LD). If found, the corresponding province field *p*_*i*_ was defined using equation 1:

**Figure 2.**
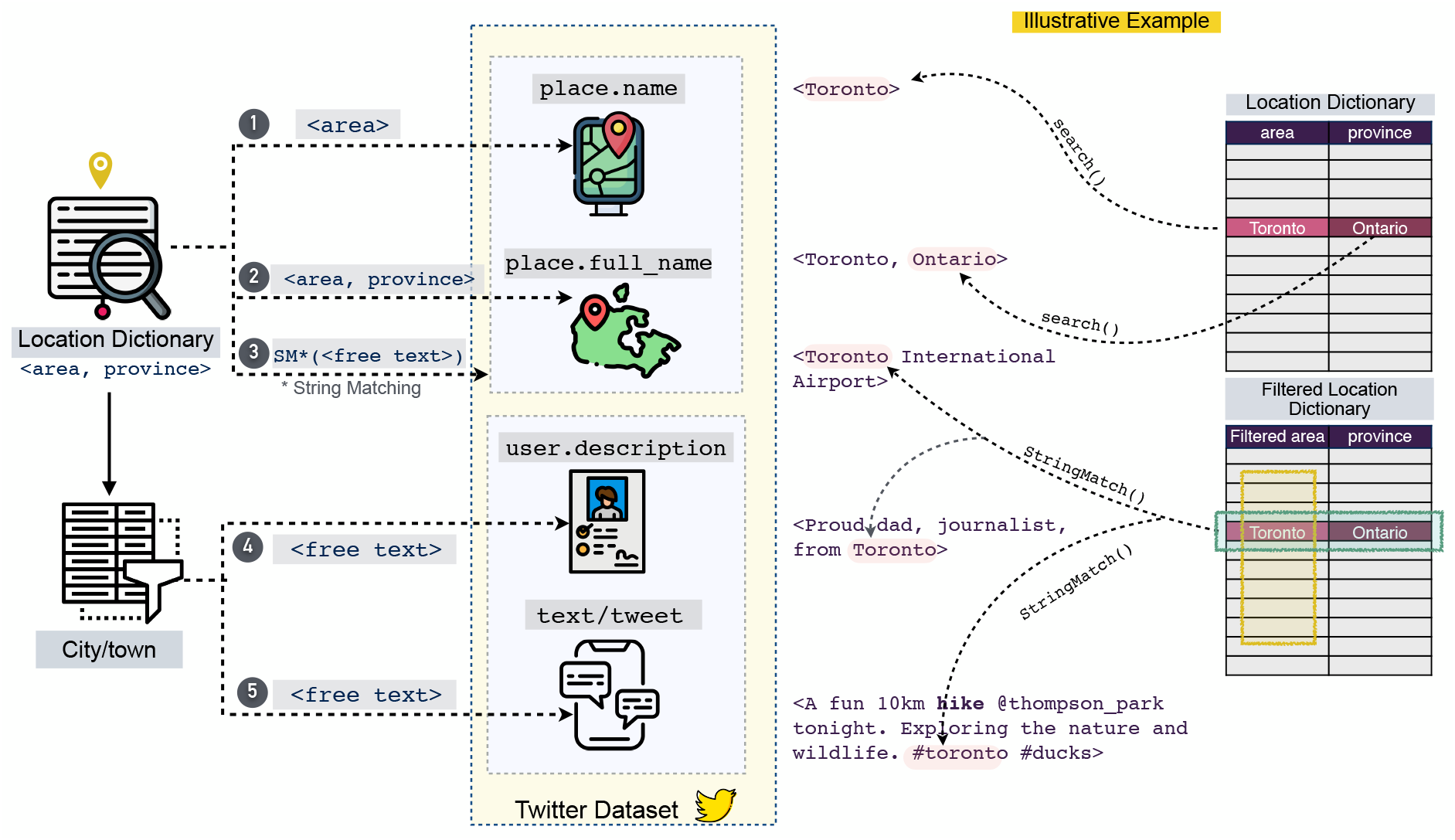
Five-step location inference process. The location dictionary is an external regional geographical metadata used to extract the exact locations of the tweets. The `area` field in this process refers to regions at different scales such as city, regional, municipality, town, township municipality, municipal district, dispersed rural community, village, or country.

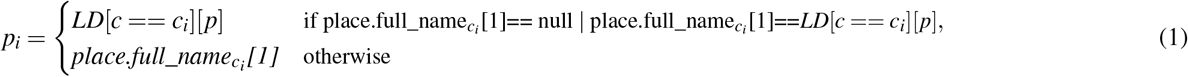

Where place.full_name_*ci*_ [1] denotes the the second component of the field when the first component is *c*_*i*_ (e.g. Ontario in the illustrative example of Figure 2). This will detect geographical areas with the same names but in different provinces (e.g. Leduc is a town/city in both Alberta and Quebec).

Steps 3-5 of the process deal with unstructured text objects that can come from all three sources of geospatial information. To extract the location information from these fields, we developed a string matching function to detect the longest common substring between the unstructured text of the dataset and the area field of the location dictionary (e.g. first time boating in Lake Louise #AB is mapped to ⟨Lake Louise, Alberta⟩ instead of to ⟨Louise, Quebec⟩). To manage the complexity of information extraction from the unstructured text, we only used a subset of areas listed in LD, with high population density (e.g. city, municipality, town, village, and country). Thus, we excluded areas classified as lake, mountain, river, bridge, or river.

### Demographic attribute inference

The demographics variable of age and gender and the information about the source of each tweet (e.g. organization vs. real users) were not available within the dataset collected from Twitter. We estimated these variables for each tweet using the `M3inference` package in `python`^26^, which uses a multimodal deep neural architecture for joint classification of age (binned into four groups: ≤18, 19-29, 30-39, ≥40), gender, and information-source of social media data. This approach uses four sources of information, including username, screen name, biography, and profile image of public profiles to develop two separate pipelines for processing a profile image and each of the three text sources of information. The models provided in this package are trained on 14.53M, 2.61M, and 23.86M profiles for each of the gender, age, and is-organisation categories, respectively.

## Data Records

LPHEADA comprises 366,405 labels for 122,135 unique tweets generated by 72,749 unique users between November 28^*th*^ 2018 to June 19^*th*^ 2020. This dataset is organized into 12 subsets (three PASS categories for each of the four countries). Table 1 provides the demographics of the dataset, including the number of tweets per PASS category for each country, labels’ characteristics, and demographics characteristics of the users. Each unique tweet is assigned a unique integer, known as `TweetID`. Each ID is mapped to the core Twitter metadata and to three crowd-generated labels for both binary and multi-class classification tasks. Figure 3 visualizes this hierarchy. As illustrated in this Figure, for each labelled tweet, LPHEADA provides the following data categories:

**Table 1.**
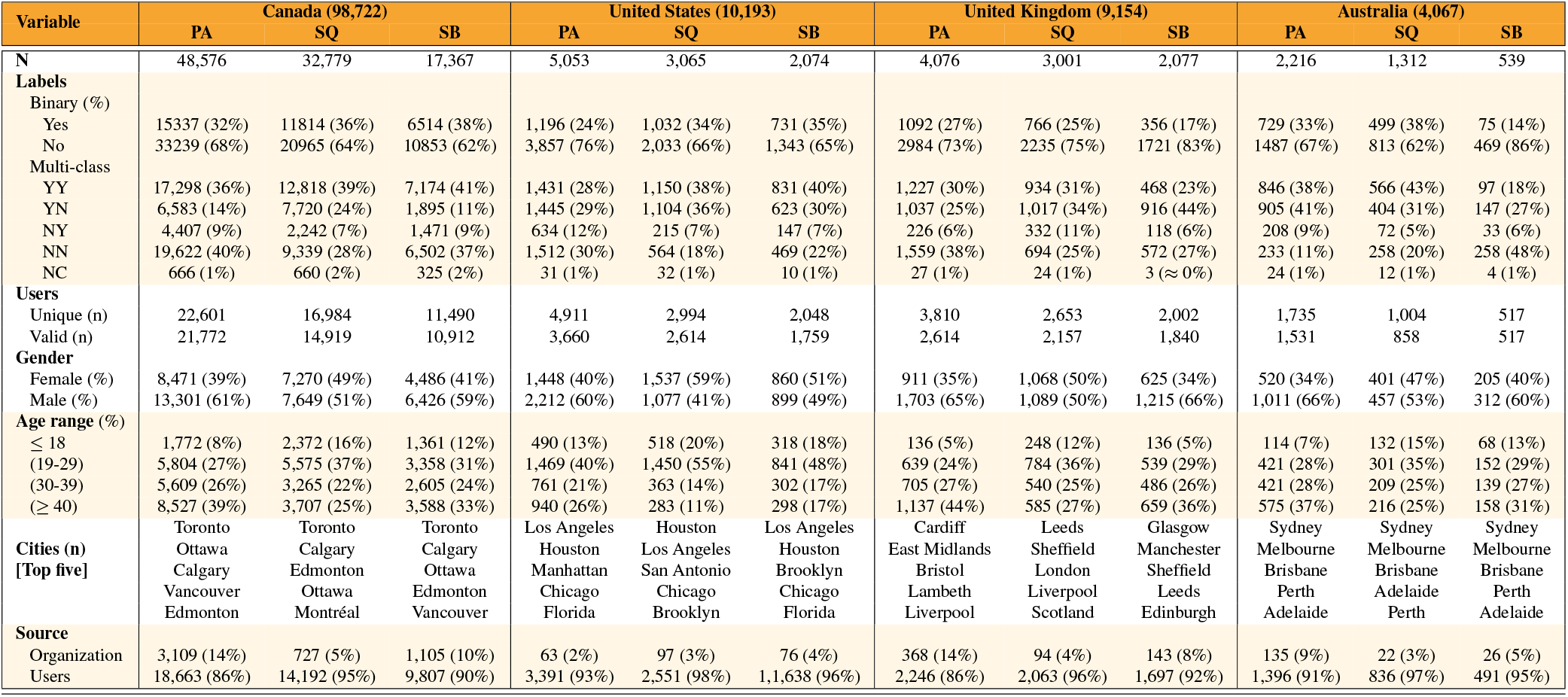
The characteristics of the dataset. The three labels collected for each tweet are consolidated into a single label using majority voting. The discrepancy between the number of binary and multi-class labels is due to the way that majority voting calculates the truth label for each of these categories.

**Figure 3.**
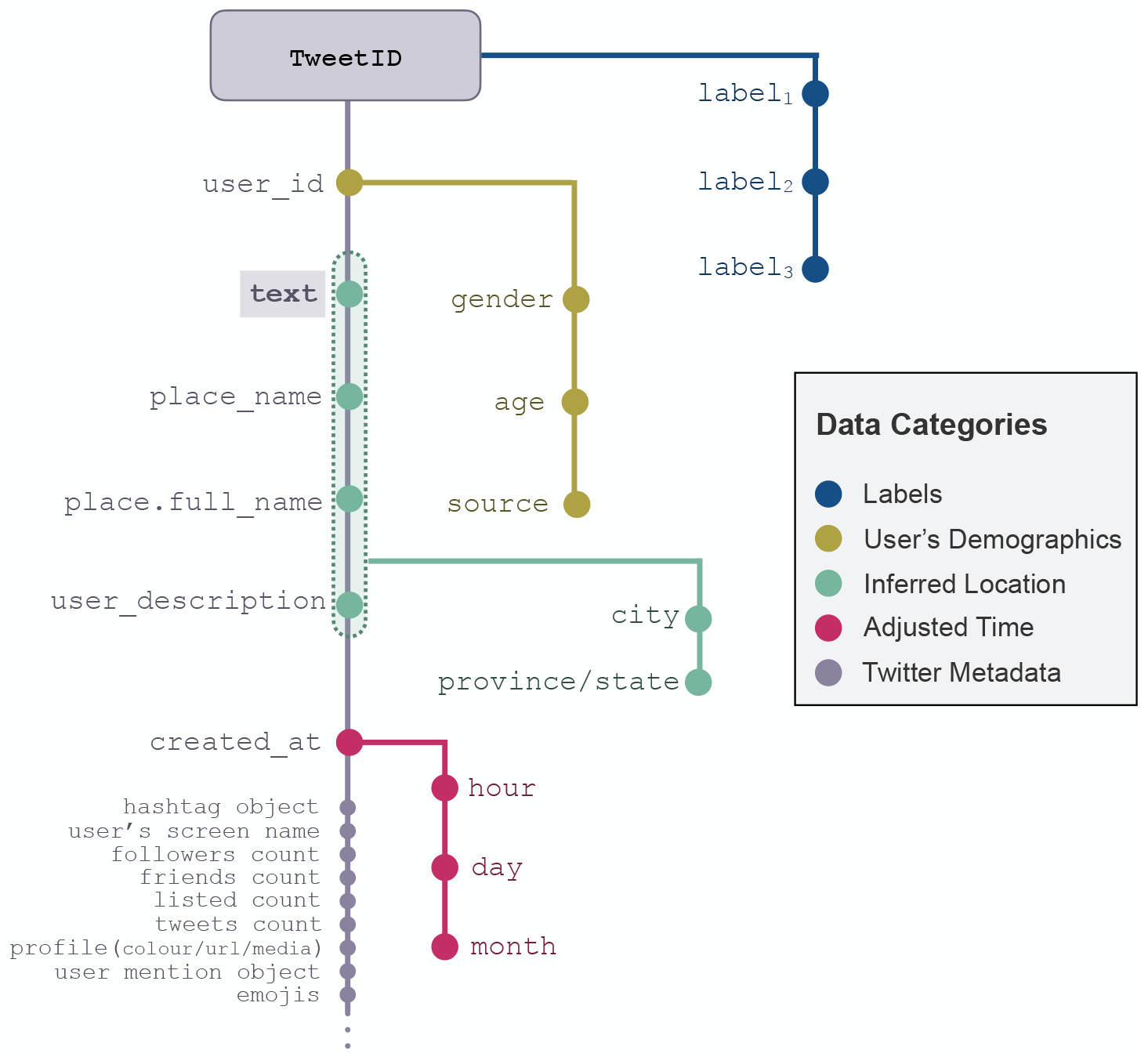
Overview of information tracking using TweetID. Each `tweet/text` is identified by a unique `TweetID` (provided by LPHEADA). This ID is mapped to metadata that include `user_id`, `place_name` `place_full_name`, `user_description`, and `created_at`. Three labels are provided for each `TweetID` that can be used for developing machine learning models. `user_id` was used to infer the demographics of each tweet including `genders`, `age` `range`, and `source`. Adjusted time (`month/day/hour`) was extracted using `created_at`, and `text`, `place_name`, `place_full_name`, and `user_description` was used to identify the `city` and `state/province` mapped to each `TweetID`.

### Labels

Let *L* denote the set of *j* unique class label, *t* present the tweet text, and *w*_*k*_ present the *k*^*th*^ worker who labeled the tweet, where *k* ∈ {1, 2, 3 }. Each *l* _*j*_∈ *L* is defined based on two conditions: whether or not the tweet is self-reported (*c*_1_ ∈ {0, 1 }); and whether or not the tweet reports a recent PASS experience (*c*_2_∈ {0, 1 }). For each PASS category across the four countries, the dataset contains the following two subsets of labels for each tweet:

#### Multi-class Labels

In this subset, each tweet *t* is mapped to quadruple ℒ = ⟨*tweetID*, (*w*_1_, *l* _*j*1_), (*w*_2_, *l* _*j*2_), (*w*_3_, *l* _*j*3_) ⟩, where *j* = 5 and each *l* _*j*_ is defined based on the value of both *c*_1_ and *c*_2_ conditions and can be formulated as {11, 10, 01, 00}. We also let workers choose a fifth option, called `Unclear`, to ensure they do not give random labels to tasks that they are not confident of performing successfully.

#### Binary Labels

Each label *l* _*j*_ in this subset is defined based on logical `AND` relationship between conditions *c*_1_ and *c*_2_. Thus, *j* = 3 with *l*_1_ = 1 if the tweet presents a self-reported PASS surveillance and *l*_1_ = 0, otherwise. Like the multi-class category, each tweet is mapped to a quadruple, and there is a class called ‘unclear’ (*l*_3_). The binary labels did not directly come from the AMT workers and were generated by dichotomizing the collected labels.

#### User’s demographics data

In the demographic dataset, each tweet *t* is mapped to quadruple *𝒟* = ⟨*tweetID, a, g, o* ⟩, where *g* ∈ {male, female} presents the gender of the user who posted the tweet, *a* ∈ {≤ 18, 19 − 29, 30 − 39, 40≤} represents their age range, and *o* ∈ {0,1} shows the source of the tweet (i.e. *o* = 1 if the tweet posted by an organization, *o* = 0 otherwise). Figure 4 shows the demographic distribution of the Canadian dataset based on gender and age-range of the unique users associated with tweet IDs. We can see that at least 75% of female users in this dataset are inferred to be younger than 40, while this number for the male users is 60%. Also, the most populated age category for female users across all PASS categories is 19-29, while this range for male users is ≥40. Excluding the (female, sleep quality) category, the age range ≤18, regardless of the user’s sex, is the least populated category across all PASS categories.

**Figure 4.**
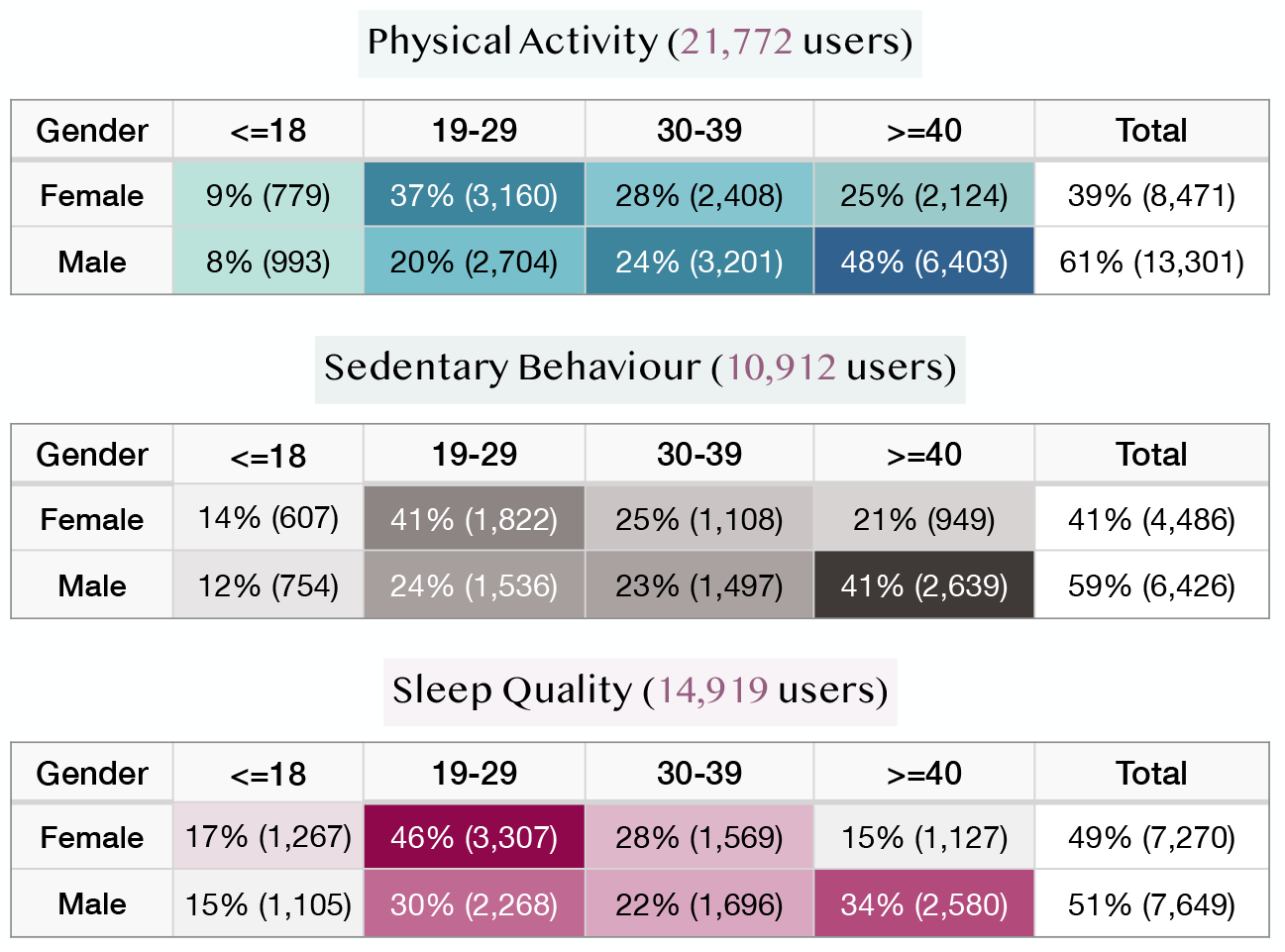
Demographic information of 42,603 unique users associated with the tweets originated in Canada.

#### Inferred location data

Each row of the location dataset is presented in the form of *𝒜* = ⟨*tweetID, c, p* ⟩, where *c* and *p* denote the city and province/state associated with each tweet, respectively. Using the TweetID parameter, the location data can be mapped to other datasets, including labels, user’s demographics, time, and twitter metadata. Each of the *c* and *p* variables are inferred based on the raw variables in Twitter metadata, including text, place objects, and user’s profile description (Figures 2 and 3). For example, Figure 5 shows the distribution of labelled tweets for each PASS category across Canadian provinces (i.e. *p*). For the top five provinces, the overall size of the dataset is directly proportional to the population size of each province. However, as only English tweets from Twitter users are included in the dataset, LPHEADA represents only English-speaking Quebecor’s and Francophone Quebecor’s tweets in English, placing the province in fourth place. Moreover, with a lower population than BC, Alberta contains more SB and SQ tweets and places in second place (Figure 5).

**Figure 5.**
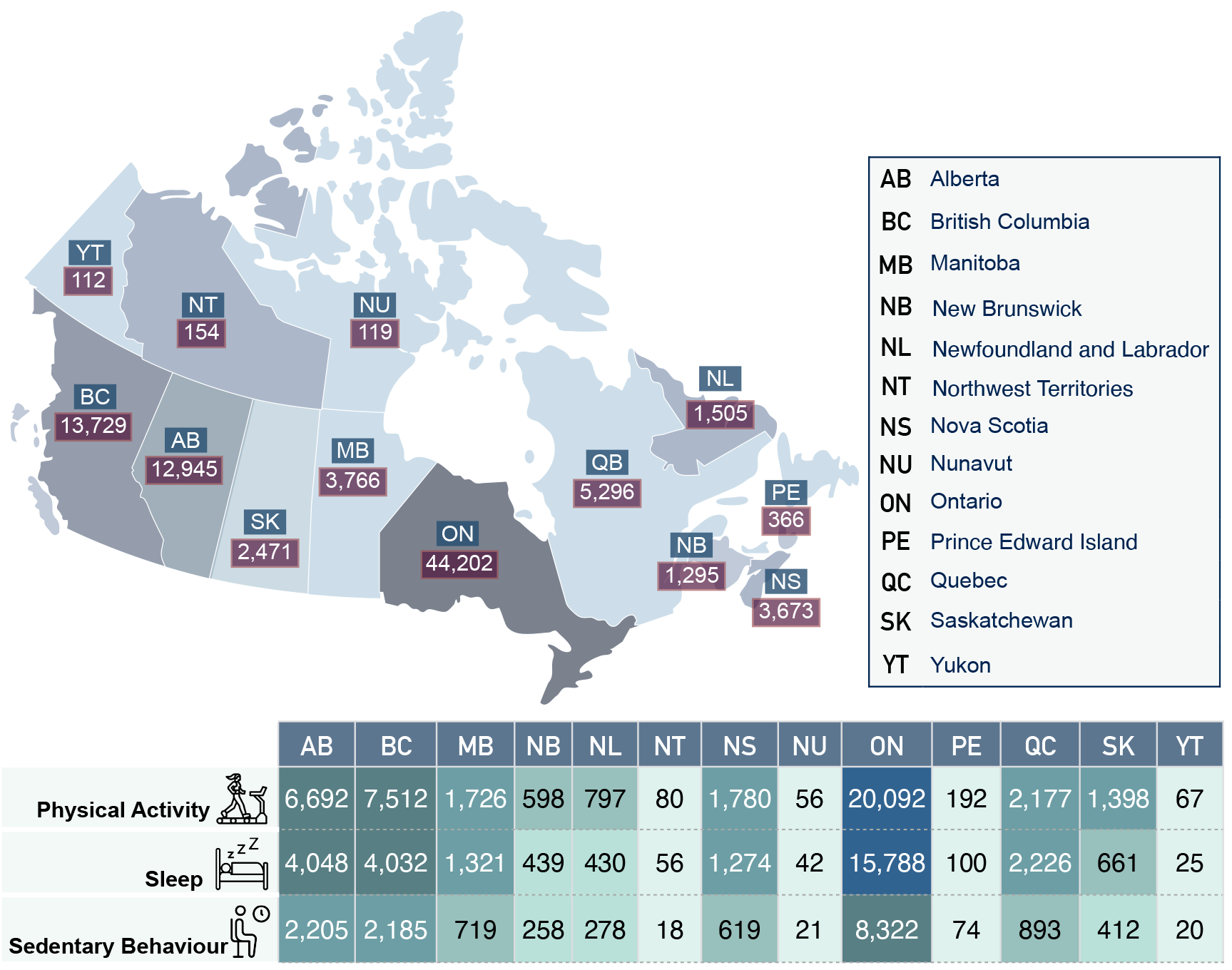
Geo-spatial details of the Canadian dataset.

#### Temporal data

The temporal dataset inferred from the ‘created_at’ field of the Twitter metadata presents the adjusted time of each tweet based on the tweet’s location. Each row of this dataset is presented in the form of *𝒯* = ⟨*tweetID, h, d, m* ⟩, where *h, d*, and *m* present the hour, weekday, and month associated with each tweet, respectively. The ‘year’ value does not need any adjustment and can be extracted directly from the original ‘created_at’ field. For example, Figure 1 represents the frequency of tweets in each PASS category across Canada at both national (Figure 1a) and city levels (Figures 1b,1c, and 1d). The highlighted area in Figure 1a demonstrates the dataset’s temporal windows that can be used to compare different aspects of PASS surveillance between 2019 and 2020.

#### Twitter metadata

In addition to the inferred data records mentioned above, TweetIDs presented in LPHEADA can be used to retrieve Twitter metadata. This metadata, in addition to the tweet text, place object, time of the tweet, and user IDs, provides more details on the tweet and user objects, including:

#### User object

This object comprises of user’s screen name, description, followers count, friends count, listed count (i.e. the number of public lists that the user is a member of), tweets count (i.e. the number of (re)tweets issued by the user), and profile characteristics (e.g. image, colour, and URLs).

#### Tweet object

This object comprises of hashtags mentioned in each tweet, emojis, user mentions, urls, and media (e.g. images and videos). For example, Figure 6 illustrates the distribution of the top 10 hashtags per label for each of the PASS categories in the entire dataset. Hashtags are basically keywords or word strings prefixed with the symbol ‘#’ that are used for categorizing and communicating tweets pertaining to the same topics. While the high level of intersection between the hashtags of positive and negative classes in our dataset makes this feature a less discriminating feature for the development of machine learning models (e.g. annotated hashtags in Figure 6), this field can still be utilized by PASS-related advocacy campaigns on Twitter to brand their movement and open up their campaigns to users who need more information about the context^27^. As tagged tweets are easily archived and accessible, the hashtag field can be effectively leveraged to improve the public’s engagement in digital PHS discussions.

**Figure 6.**
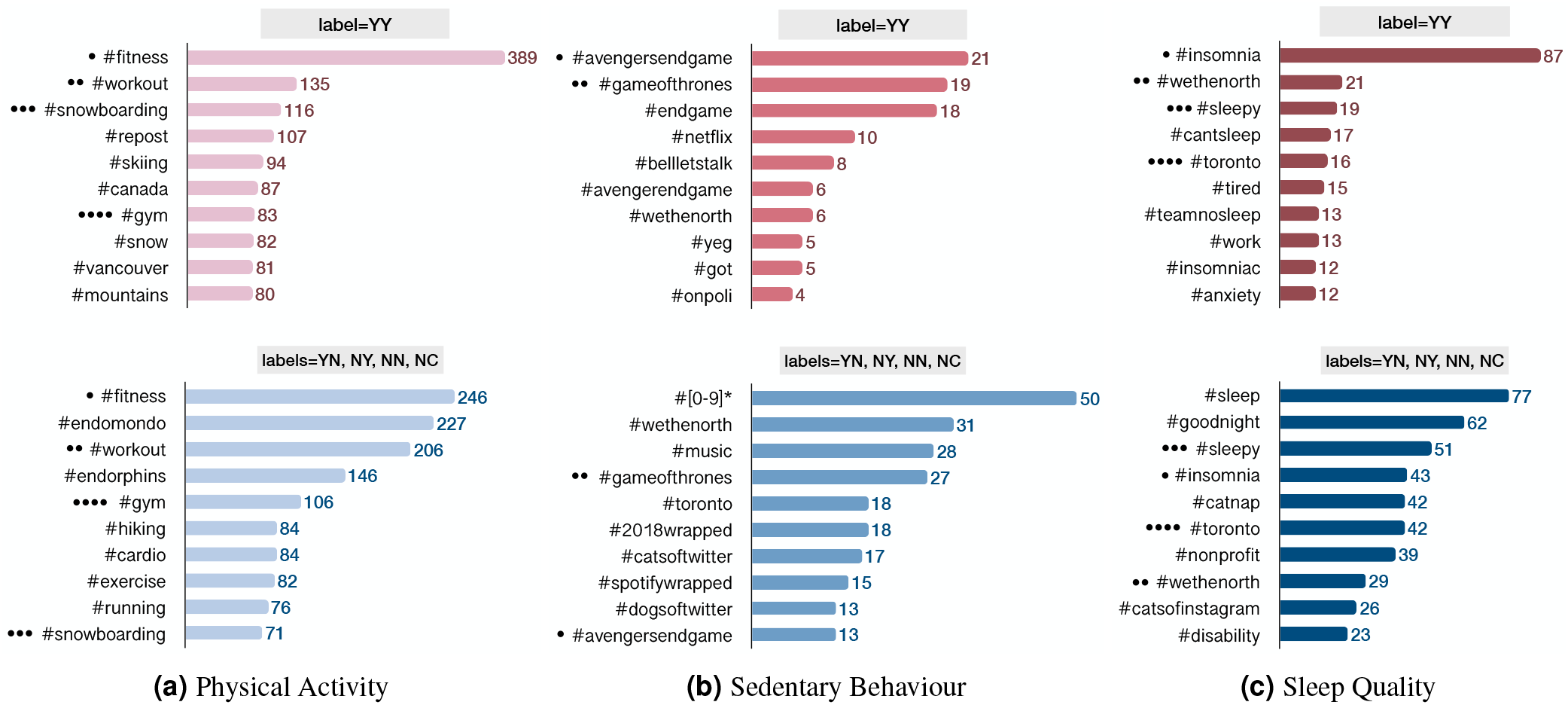
The distribution of top 10 hashtags per label for each of the physical activity, sedentary behaviour, and sleep quality categories. The number at the end of each bar presents the frequency of its corresponding hashtag. The intersections between two class of labels for each PASS category are annotated using filled circles (•). This Figure is based on all data collected from Canada, US, UK, and Australia.

## Technical Validation

To verify the quality of crowd-generated labels and set a baseline for the dataset, we conducted four studies. First, we used a series of statistical inference models to verify the quality of the labels provided in this dataset. Second, we evaluated the semantic consistency between the datasets collected from the countries included in our repository. Third, we trained and tested 12 binary classifiers using labels provided in the dataset. Finally, to investigate the structural differences between all subsets of LPHEADA, we conducted linguistic and lexical analysis and visualized the results for further comparisons. Moreover, to address unseen technical issues of the dataset, we provide a public issue tracker for handling bug reports, describing solutions to technical issues, data updates, and other issues and contributions.

### Methods of label agreement

To measure the consistency of labels generated by AMT workers, we calculated label consistency (ℒ *C*) as the average entropy of the collected labels for each PASS category^28^. For each tweet *t*_*i*_ ∈ *𝒯* _*s*_, where *𝒯* _*s*_ denotes the set of all tweets related to surveillance category *s* ∈ {physical activity, sleep quality, sedentary behaviour}, *n*_*i j*_ defines the number of answers given to the *j*^*th*^ choice (*j* ∈ {1, 2, 3, 4, 5}), as we have five choices for each tweet). We calculate *ℒC*_*s*_ as:

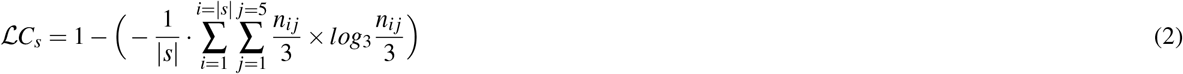

|*s*| denotes the size of the surveillance category *s* and as we collect three labels for each tweet, the denominators in the entropy formula receive the constant value of three. *ℒC* ranges from 0 to 1, and the values close to zero show less consistency between the workers’ input. After calculating the label consistency (*ℒC*) for each PASS category, we had *ℒC >* 0.52 for the multi-class labelling, and *ℒC >* 0.73 for the binary labelling tasks.

To consolidate the collected labels for each tweet, we used the majority voting (MV) technique (Table 1). Defining the estimated label as 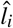, and the submitted label by worker *w* as *l*_*w*_, the MV approach, for a binary labeling task, assigns 1 to 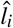 if 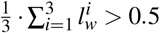, and assigns 0, otherwise. The discrepancy between the number of binary and multi-class labels presented in Table 1 caused by the way that MV approach calculates the truth label for each of these categories. In addition to MV, there are models, such as David and Skene (DS)^29^, Raykar (RY)^30^, and generative model of labels, abilities, and difficulties (GLAD)^31^, that incorporate the error rate of annotators (workers), task complexity, and context-sensitive features into the inference process and can be utilized to predict truth labels from crowd-labelled data.

### Semantic consistency

To validate the semantic consistency of the datasets collected from different countries, we transformed the dataset of each PASS category into a semantic space of low dimensionality using latent semantic analysis (LSA). For the vector presentation of each dataset, to capture high-level semantics of the text, we ran the PASS category of each country through a pre-trained word2vec embedding model. This model contains 300-dimensional vectors of three million words and phrases trained on 100 billion words from a Google News dataset. Then, the resulting 300-dimensional vectors were averaged for each tweet. For each tweet

*T* composed of words ⟨*w*_1_, *w*_2_, …, *w*_*n*_⟩, with 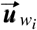. defining the embedding of *w*_*i*_, the embedding of tweet *T* can be calculated as: 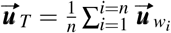 We then applied truncated SVD on the new vectorized dataset and only kept the top two dimensions of the dataset containing the most variance (e.g. those directions in vector space of the dataset that contain more information). The scatter plots presented in Figure 7 illustrate our datasets in a two-dimensional latent semantic space. The high level of overlap between the datasets of each PASS category indicates that the data from different countries cover similar semantic space, but the space is scaled differently based on the size of datasets.

**Figure 7.**
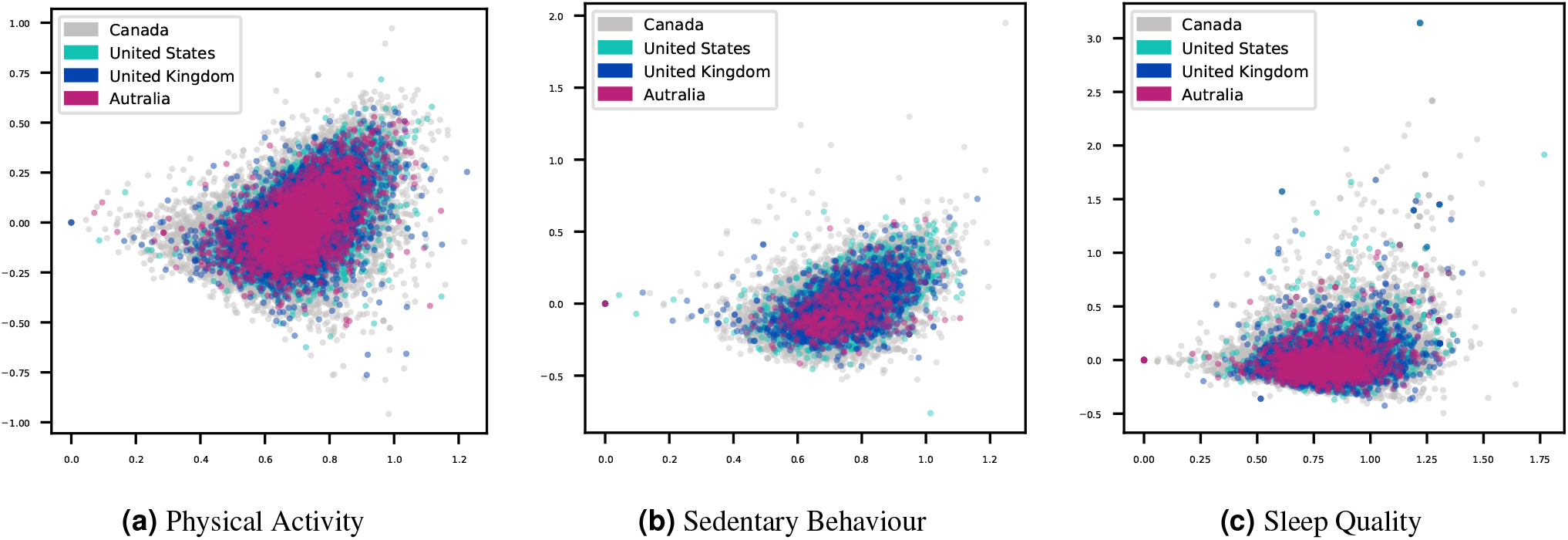
Scatter plots of the first two dimensions of latent semantic analysis (LSA) performed on PASS categories and classified based on the geographic source of the data.

Moreover, to further investigate the internal consistency of the datasets presented in this paper, we trained three convolutional neural network (CNN) multiclass classifiers (i.e. one for each PASS category) to classify the tweets into one of Canada, USA, Australia, or UK classes. Given the highly imbalanced distribution of the classes in our dataset due to the highly unequal number of samples from each country, we used average precision metric (AP) to measure the discrimination ability of our predictive models. The poor performance of these classifiers (AP_*PA*_: 37%, AP_*SB*_: 32%, AP_*SQ*_: 31%) in detecting each tweet’s country implies a high level of semantic and syntactic cohesion among the 4 countries in our dataset.

### Classification of PASS categories

For the PASS classification experiment, we utilized a standard CNN classifier with one layer of convolution with global max-pooling on top of a word2vec embedding trained on 100 billion words of Google News. The vectors have a dimensionality of 300 and were trained using the continuous bag-of-words architecture^32^. We used the binary labels of the dataset to train and evaluate the model on each of 12 datasets provided in LPHEADA. Due to the imbalanced distribution of binary labels across all of these datasets (Table 1), in addition to precision, recall, F1, and AUC scores, we used average precision (AP) to measure the weighted mean of precision at different thresholds to make the score robust to heterogeneous and imbalanced class distributions. Like AUC score, AP is a model-wide and threshold-free evaluation metric. However, for imbalance class distributions with the negatives outnumbering the positives, AP is more informative than AUC, as it mainly evaluates the fraction of true positive samples among positive predictions and is more robust to the relationship between false-positive (FP) and false-negative (FN) rates^33^. As shown in Table 2, for each of the Canada, USA, and UK datasets, we find a steady increase in the overall performance of the classifier as the size of the dataset increases (i.e. | *PA* |*>*| *SQ* |*>*| *SB* |). Interestingly, the UK dataset achieves the highest performance for the PA category than all the other countries.

**Table 2.** Binary classification of tweet using bidirectional LSTM. We used the same classifier to classify the dataset based on the countries: AP (PA: 37%, SB: 32%, and SQ: 31%).

| Physical Activity |  |  |  |  |
| --- | --- | --- | --- | --- |
| Metrics | Canada | United States | United Kingdom | Australia |
| Precision | 78% | 75% | 80% | 65% |
| Recall | 78% | 76% | 81% | 68% |
| F1 | 78% | 76% | 80% | 64% |
| AUC<sub>ROC</sub> | 83% | 76% | 84% | 64% |
| AP | 81% | 75% | 81% | 66% |
| Sedentary Behaviour |  |  |  |  |
| Metrics | Canada | United States | United Kingdom | Australia |
| Precision | 73% | 62% | 79% | 74% |
| Recall | 73% | 64% | 83% | 86% |
| F1 | 73% | 62% | 79% | 80% |
| AUC<sub>ROC</sub> | 76% | 62% | 63% | 57% |
| AP | 78% | 65% | 66% | 66% |
| Sleep Quality |  |  |  |  |
| Metrics | Canada | United States | United Kingdom | Australia |
| Precision | 76% | 70% | 73% | 61% |
| Recall | 76% | 70% | 76% | 60% |
| F1 | 75% | 70% | 73% | 60% |
| AUC<sub>ROC</sub> | 81% | 70% | 66% | 63% |
| AP | 83% | 72% | 71% | 64% |

### Linguistic Properties

To understand and validate the linguistic properties of each dataset, we measured and visually compared the following metrics for each PASS category grouped by countries: (i) sentence count, (2) grammar score (i.e. number of grammar errors), (3) the average number of syllables per word and the average sentence length (i.e. Flesch-Kincaid Grade Level index^34^), (4) the average number of words per sentence and the percentage of words with three or more syllables (i.e. Gunning Fog index^35^), (5) a combination of average sentence length and percentage of difficult words (i.e. Dale–Chall readability^36^), (6) Sentence length and number of polysyllables (i.e. Linsear Write readability^36^), (7) number of characters (i.e. Coleman-Liau Index^36,37^), (8) the average number of characters per word and number of words per sentence (i.e. automated readability index^38^), and (9) the text standard score based on number of sentences, words, syllables, and characters in each tweet (i.e. text readability consensus). Figure 8 illustrates these comparisons based on the minimum (red), average (pink), and maximum (light pink) value of each feature. While all datasets have similar behaviour in terms of each feature’s minimum value, the Canadian dataset has a lower score for the average number of syllables per word and the average sentence length for all PASS categories. Interestingly, the sleep quality dataset, compared to other PASS categories, has a higher value for the maximum number of ‘grammar errors’ and ‘sentence count’ metrics, while all datasets show the same behaviour for the minimum and the average values of these metrics. These location-specific linguistic characteristics should be considered when utilizing these datasets to train and evaluate PASS surveillance ML models. For example, a model trained on the Canadian dataset may not present some linguistic features of a dataset that originated in Australia and vice versa.

**Figure 8.**
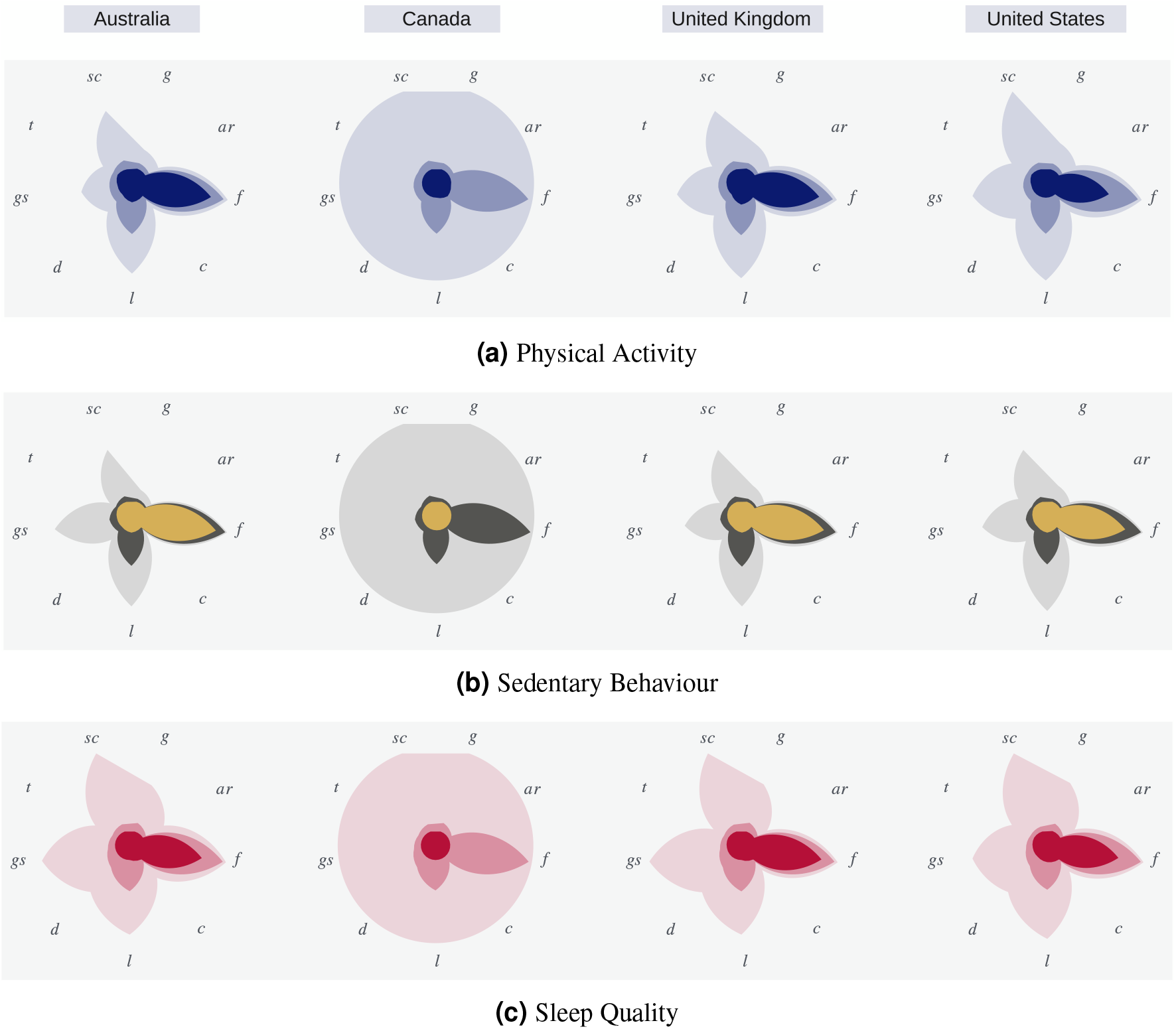
*sc*: sentence count, *g*: Gunning Fog index, *ar*: automated readability index, *f* :Flesch-Kincaid grade level, *c*: Coleman-Liau Index, *l*: Linsear Write readability, *d*: Dale–Chall readability, *gs*: grammar score, and *t*: text readability consensus.

## Usage Notes

### Data access

LPHEADA is publicly available online. Retrieving the data using TweetIDs provided in the dataset requires Twitter’s approval. Data access also requires a data use agreement between the data user and Twitter to govern the access and use of the licensed material returned by Twitter API. Once approved, Twitter metadata can be downloaded as a JSON file to be mapped to other subsets of data (e.g. location, time, and demographics) using the TweetID field.

## Example Usage

We have provided publicly accessible instructions and Jupyter Notebooks^39,40^ to illustrate the application of the data (available at https://github.com/data-intelligence-for-health-lab/Lpheada-Labelled-Public-HEAlth-DAtaset).

## Code availability

All code used in the experiments described in the manuscript was written in Python 3 and is available through our GitHub repository (https://github.com/data-intelligence-for-health-lab/Lpheada-Labelled-Public-HEAlth-DAtaset). We provide all necessary instructions, required libraries, and sample Jupyter Notebooks allowing replicating our experiments and utilizing the dataset.

## Acknowledgements

This work was supported by a postdoctoral scholarship from the Libin Cardiovascular Institute and the Cumming School of Medicine, University of Calgary. Also, this work was supported by a Discovery Grant from the Natural Sciences and Engineering Research Council of Canada (RGPIN-2014-04743). The Public Health Agency of Canada funded the Amazon Mechanical Turk costs.

## Author contributions statement

Z.SH. was responsible for data collection and curation, model development, documentation and maintenance of the dataset, data analysis and visualization and wrote the paper. G.B. and W.T. reviewed the paper and provided comments. J.L. contributed to the conception and design of the study and revised the manuscript.

